# VIDEO BASED DETECTION OF EPILEPTIC SEIZURES USING A THREE-DIMENSIONAL CONVOLUTIONAL NEURAL NETWORK

**DOI:** 10.1101/2024.10.11.24315247

**Authors:** Aidan Boyne, Hsiang J. Yeh, Anthony K. Allam, Brandon M. Brown, Mohammad Tabaeizadeh, John M. Stern, R. James Cotton, Zulfi Haneef

## Abstract

**Objective:** Seizure detection in epilepsy monitoring units (EMU) is essential for the clinical assessment of drug-resistant epilepsy. Automated video analysis using machine learning provides a promising aid for seizure detection with resultant reduction in the resources required for diagnostic monitoring. We employ a 3D convolutional neural network with fully fine-tuned backbone layers to identify seizures from EMU videos.

**Methods:** A two-stream inflated 3D-ConvNet architecture (I3D) classified video clips as a seizure or not a seizure. A pretrained action classification model was fine-tuned on 11 hours of video data containing 49 tonic-clonic seizures from 25 patients monitored at a large academic hospital (site A) using leave-one-patient-out cross-validation. Performance was evaluated by comparing model predictions to ground-truth annotations obtained from video-EEG review by an epileptologist on videos from site A and a separate dataset from a second large academic hospital (site B).

**Results:** The model achieved leave-one subject out cross-validation F1-score of 0.960 ± 0.007 and area under the receiver operating curve (AUC) score of 0.988 ± 0.004 at site A. Evaluation on full videos successfully detected all seizures with median detection latency of 0.0 (0.0, 3.0) seconds from seizure onset. The site A model had an average false alarm rate of 1.81 alarms per hour, though 33 of the 49 videos (67%) had no false alarms. Evaluation at site B demonstrated generalizability of the model architecture and training strategy, though cross-site evaluation (site A model tested on site B data and vice versa) resulted in diminished performance.

**Significance:** Our model demonstrates high performance in the detection of epileptic seizures from video data using a fine-tuned I3D model and outperforms prior similar models identified in the literature. This study provides a foundation for future work in real-time EMU seizure monitoring and possibly for reliable and cost-effective at-home detection of tonic-clonic seizures.

**KEY POINTS:**

- We evaluate a video-based 3-D CNN for seizure detection in patients undergoing evaluation in an EMU at 2 large academic hospitals.
- Our video-only model provides highly accurate detection of tonic-clonic seizures with low detection latency.
- The underlying model architecture requires no video preprocessing and is generalizable across two EMUs.

## 1. INTRODUCTION

Accurate and timely detection of epileptic seizures is critical for safe and efficient video-EEG evaluations and would improve patient safety at home when alone (1). Evaluation in an epilepsy monitoring unit (EMU) is the standard evaluation for drug-resistant epilepsy and requires continuous monitoring by a care provider for safety. However, EMU access is often insufficient for the care needs of a community, and EMU admission is both expensive and time intensive for hospital staff (2). Alternatives to the EMU, such as wearable multimodal seizure detection devices using accelerometers and surface electromyography, have shown promise but have high false alarm rates and are often impractical for prolonged use (3,4).

Recent advances in computer vision and deep learning offer a potential solution to these limitations by providing non-contact seizure detection via video data analysis. Automated detection of seizures based only on video would allow at-home seizure monitoring and alert systems. Accurate and precise detection software could form the basis of at-home and inpatient seizure alarms and play a role in reducing sudden unexpected death in epilepsy (5).

The numerous advantages of video-based seizure detection have made the subject an area of active research. Detection of the characteristically abnormal movement and/or posturing during seizures underlies the majority of preexisting work. Older methods primarily relied on tracking key points of interest, such as the joints and eyes, or measuring the amplitude and direction of limb and body movement (6–8). Performance of these models was generally low and was likely limited by machine learning techniques such as support vector machines that could utilize only a tiny fraction of the data available in the video recording. However, deep learning allows researchers to take advantage of the larger amount of information available in the video data.

Many models using convolutional neural networks (CNN) have produced impressive results in seizure detection and classification tasks using a wide range of input data types (9–13) and have largely outperformed other model architectures (14). In particular, work by Karácsony et al. demonstrated the potential of 3D classifiers as feature extractors in seizure classification pipelines (9). In this investigation, we present an approach that uses a 3D-CNN-based algorithm with fine-tuned backbone layers and classification head for the automated detection of seizures from spatiotemporal video input.

## 2. METHODS

### 2.1 Data Acquisition

All data were acquired from patients undergoing long-term video-EEG monitoring in the EMU at the Baylor St. Luke’s Medical Center. The study was approved by the Institutional Review Board at Baylor College of Medicine under protocol H-47804. Videos were recorded at 30 frames per second at a resolution of 640×480 pixels using a SONY SNC EP520 camera mounted on the ceiling of the EMU patient room. These cameras automatically acquire color (RGB) video during high light conditions and greyscale video in low light conditions. Audio was also recorded but was not used in training or testing of the model. Videos corresponding to electro-clinical seizures underwent manual review by experts for visual evidence of generalized and focal seizures containing visible tonic-clonic and were segmented into seizure and non-seizure sections accordingly. Videos without any visually identifiable tonic-clonic seizures were excluded from the analysis. Two additional videos were excluded from the final analysis based on the patient being out of the video’s frame, overly obstructed, or out of focus. EEG readings were captured using conventional scalp EEG montages on a Nihon Kohden system using the international 10-20 system of electrode placement. EEG data were used only for labeling the videos and were not used in the training or testing of the model.

### 2.2 Dataset Preparation

A total of 21 hours of video data was collected from 25 patients. Clinical annotations were used to identify EMU videos for potential inclusion and sections of the video containing any visually evident seizure activity were marked. After excluding monitoring sessions without movement, 49 videos from the 25 patients remained, totaling 11 hours and 4 minutes of usable video data. This data consisted of 55 minutes of seizure activity (8.3%) and 10 hours and 9 minutes (91.7%) of non-seizure activity based on the manual review described in section 2.1. (**Table 1**). The data were then divided into 3-second-long clips for model training and validation. Clips were labeled as seizure if the clip contained at least one second of seizure-related movements.

**Table 1:** Breakdown of EMU monitoring video footage by patient and event. Each event is captured and converted to a .mp4video file. Fellowship trained epileptologists reviewed each video along with the corresponding EEG trace to classify segments of the video as seizure or non-seizure. Abbreviations: GS, Greyscale; RGB, Red Green Blue.

| Patient | Video ID | Seizure Length (s) | Seizure % | Non-Seizure Length (s) | Non-Seizure % | Total Length (s) | Color |
| --- | --- | --- | --- | --- | --- | --- | --- |
| **1** | P1-V1 | 59 | 8.76% | 614 | 91.24% | 673 | GS |
|  | P1-V2 | 62 | 9.27% | 607 | 90.73% | 669 | RGB |
|  | P1-V3 | 58 | 5.21% | 1055 | 94.79% | 1113 | RGB |
| **2** | P2-V1 | 78 | 6.18% | 1185 | 93.82% | 1263 | RGB |
| **3** | P3-V1 | 94 | 11.80% | 703 | 88.20% | 797 | RGB |
| **4** | P4-V1 | 70 | 7.82% | 826 | 92.18% | 896 | RGB |
|  | P4-V2 | 80 | 5.96% | 1263 | 94.04% | 1343 | RGB |
|  | P4-V3 | 89 | 13.15% | 588 | 86.85% | 677 | RGB |
| **5** | P5-V1 | 79 | 11.83% | 589 | 88.17% | 668 | GS |
| **6** | P6-V1 | 57 | 3.45% | 1595 | 96.55% | 1652 | RGB |
| **7** | P7-V1 | 78 | 6.83% | 1064 | 93.17% | 1142 | GS |
| **8** | P8-V1 | 50 | 5.74% | 821 | 94.26% | 871 | RGB |
| **9** | P9-V1 | 81 | 21.80% | 291 | 78.20% | 372 | RGB |
| **10** | P10-V1 | 75 | 20.53% | 290 | 79.47% | 365 | RGB |
| **11** | P11-V1 | 34 | 10.58% | 287 | 89.42% | 321 | GS |
|  | P11-V2 | 44 | 9.97% | 397 | 90.03% | 441 | RGB |
|  | P11-V3 | 31 | 10.20% | 273 | 89.80% | 304 | GS |
|  | P11-V4 | 39 | 11.24% | 308 | 88.76% | 347 | RGB |
|  | P11-V5 | 38 | 8.61% | 403 | 91.39% | 441 | RGB |
|  | P11-V6 | 40 | 13.55% | 255 | 86.45% | 295 | RGB |
| **12** | P12-V1 | 62 | 14.34% | 370 | 85.66% | 432 | RGB |
| **13** | P13-V1 | 65 | 6.34% | 961 | 93.66% | 1026 | RGB |
| **14** | P14-V1 | 66 | 6.42% | 962 | 93.58% | 1028 | GS |
|  | P14-V2 | 75 | 9.79% | 691 | 90.21% | 766 | GS |
|  | P14-V3 | 69 | 16.35% | 353 | 83.65% | 422 | RGB |
|  | P14-V4 | 67 | 5.90% | 1068 | 94.10% | 1135 | GS |
| **15** | P15-V1 | 75 | 3.94% | 1831 | 96.06% | 1906 | RGB |
| **16** | P16-V1 | 65 | 5.87% | 1042 | 94.13% | 1107 | RGB |
| **17** | P17-V1 | 80 | 16.44% | 407 | 83.56% | 487 | RGB |
|  | P17-V2 | 68 | 8.33% | 748 | 91.67% | 816 | RGB |
|  | P17-V3 | 84 | 4.45% | 1806 | 95.55% | 1890 | RGB |
|  | P17-V4 | 69 | 30.45% | 158 | 69.55% | 227 | RGB |
|  | P17-V5 | 73 | 19.99% | 292 | 80.01% | 365 | GS |
|  | P17-V6 | 48 | 25.11% | 143 | 74.89% | 191 | RGB |
| **18** | P18-V1 | 43 | 4.69% | 874 | 95.31% | 917 | RGB |
| **19** | P19-V1 | 98 | 8.13% | 1108 | 91.87% | 1206 | RGB |
|  | P19-V2 | 125 | 9.02% | 1261 | 90.98% | 1386 | GS |
| **20** | P20-V1 | 66 | 11.13% | 527 | 88.87% | 593 | RGB |
|  | P20-V2 | 44 | 6.31% | 653 | 93.69% | 697 | RGB |
| **21** | P21-V1 | 198 | 12.21% | 1424 | 87.79% | 1622 | RGB |
| **22** | P22-V1 | 65 | 6.36% | 957 | 93.64% | 1022 | RGB |
| **23** | P23-V1 | 70 | 7.11% | 915 | 92.89% | 985 | RGB |
|  | P23-V2 | 61 | 9.00% | 617 | 91.00% | 678 | RGB |
|  | P23-V3 | 48 | 5.46% | 831 | 94.54% | 879 | RGB |
|  | P23-V4 | 62 | 10.37% | 536 | 89.63% | 598 | RGB |
| **24** | P24-V1 | 61 | 5.58% | 1032 | 94.42% | 1093 | GS |
| **25** | P25-V1 | 41 | 6.04% | 638 | 93.96% | 679 | RGB |
|  | P25-V2 | 57 | 11.01% | 460 | 88.99% | 517 | RGB |
|  | P25-V3 | 52 | 9.22% | 512 | 90.78% | 564 | RGB |
| **Total** |  | **3293** | **8.26%** | **36589** | **91.74%** | **39882** |  |

### 2.3 Model Structure

A two-stream inflated 3D CNN (I3D) was used as the base model for video classification (**Figure 1**). This model architecture demonstrated excellent results in human action classification and achieved promising results when used for feature extraction in the classification of frontal vs. temporal lobe epilepsy (9). The architecture is described in detail in the seminal work by Carreira and Zisserman (15). The structure of the model in the context of seizure detection is briefly described here.

**Figure 1:**
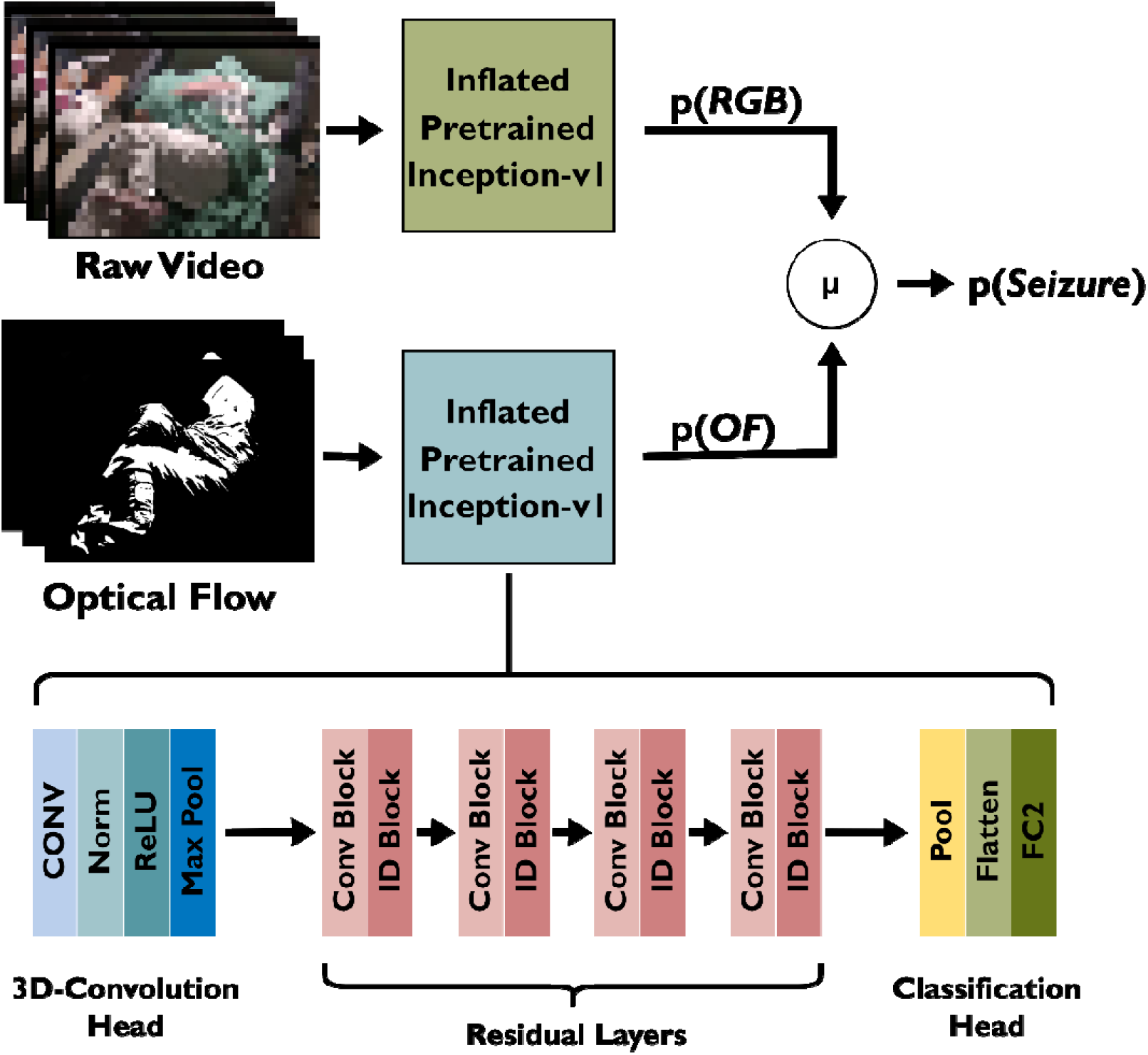
The I3D model consists of two inflated Inception-v1 sub-models. These sub-models were fine-tuned on Image Net Data and the entire I3D model was then fine-tuned on the Kinetics Human Action Video 400 dataset *(16,17)*. The 3D-convolutional head extracts spatiotemporal features, stabilizes the input distribution, and introduces non-linearity to the model before reducing the size of the input and passing it to the residual layers. These layers are defined by residual connections that allow the model to skip certain layers, leading to a more stable and convergent end result. Finally, the output of the residual layer is passed to the classification head which outputs a probability from 0 (not seizure) to 1 (seizure). Three-second video clips taken from the EMU were then fed to the model training pipeline which automatically calculates OF for each clip before sending the OF and RGB videos to the appropriate submodel. (Abbreviations: RGB, red-green-blue; p(RGB), Raw video stream predicted seizure probability; p(OF), optical flow stream predicted seizure probability; µ, arithmetic mean; p(Seizure); overall model predicted seizure probability; CONV, convolutional layer; Norm, normalization layer; ReLU, rectified linear unit activation function; ID Block, identity block; FC2, fully connected 2 class layer.)

The model consists of an initial 3D convolution layer followed by 2 spatiotemporal max-pooling layers. The 3D convolution layers allow the model to capture both spatial (image edges, textures, and shapes) and temporal (change in frames over time) features from the EMU videos. Output from the 3D convolution is compressed by the pooling layers and fed through four residual layers composed of multiple Bottleneck3D blocks which progressively reduce spatial dimensions while increasing the depth of the feature maps. Finally, the features are used as input for the classification head. The original classification head designed for the recognition of 400 actions was reduced from 400 to 2 outputs to allow for binary (seizure or non-seizure) classification of videos. The modified classification head outputs a probability ranging from 0 to 1 where 0 represents no likelihood of seizure activity and 1 represents certain seizure activity.

The two streams of data used for classification were a raw RGB or greyscale video and the same video fed through an optical flow algorithm. The raw video stream provided spatial features such as objects and textures to the model, while the optical flow stream captured motion between consecutive frames of the video. The predictions of each stream were averaged to provide the final predicted probability of seizure activity within the video (**Figure 1**). Each video clip fed to the model was classified as seizure if the final predicted probability was greater than 0.5.

### 2.4 Model Training and Evaluation

Two identical 2D-Inception-v1 models pretrained on ImageNet (17), a gold standard image dataset for object recognition tasks, were used as the backbone of our seizure detection model. Weights from the models were inflated to allow the models to process spatiotemporal (3D) video data. The inflation process copies the initial model’s 2D kernels multiple times along a third axis to make 3D kernels, analogous to stacking multiple 2D squares on top of one another to form a 3D cube. The temporally inflated Inception-v1 models were then fine-tuned using data from the Kinetics Human Action Video 400 dataset, a dataset of various labeled videos often used to train models to recognize human activities, resulting in a general-purpose action recognition I3D model (16). This model is publicly available and has demonstrated high performance on action recognition tasks (15,18). Lastly, the output layer of the individual Inception-v1 sub-models was shrunk from 400 to 2 outputs to allow for binary (seizure or non-seizure) classification of videos.

We used the MMaction2 training framework with a cross-entropy loss function to fine tune the weights of both the model backbone and classification head on our seizure dataset (19). The framework is optimized for training and testing human action recognition models and was well suited for fine-tuning the I3D model for seizure detection. All training and validation were performed using a GeFORCE RTX 4090 graphics card [NVIDIA Corporation, Santa Clara, CA].

The models were trained and evaluated using leave-one-patient-out (LOPO) cross-validation to prevent data leakage. For each fold, all video clips from one patient were set aside as a test set. All seizure video clips and an equal number of randomly selected non-seizure video clips from the remaining patients were then combined to form a balanced dataset. The balanced dataset was into training and validation sets at a ratio of 80:20. To evaluate the model, both balanced and unbalanced test sets were used (**Figure 2**).

**Figure 2:**
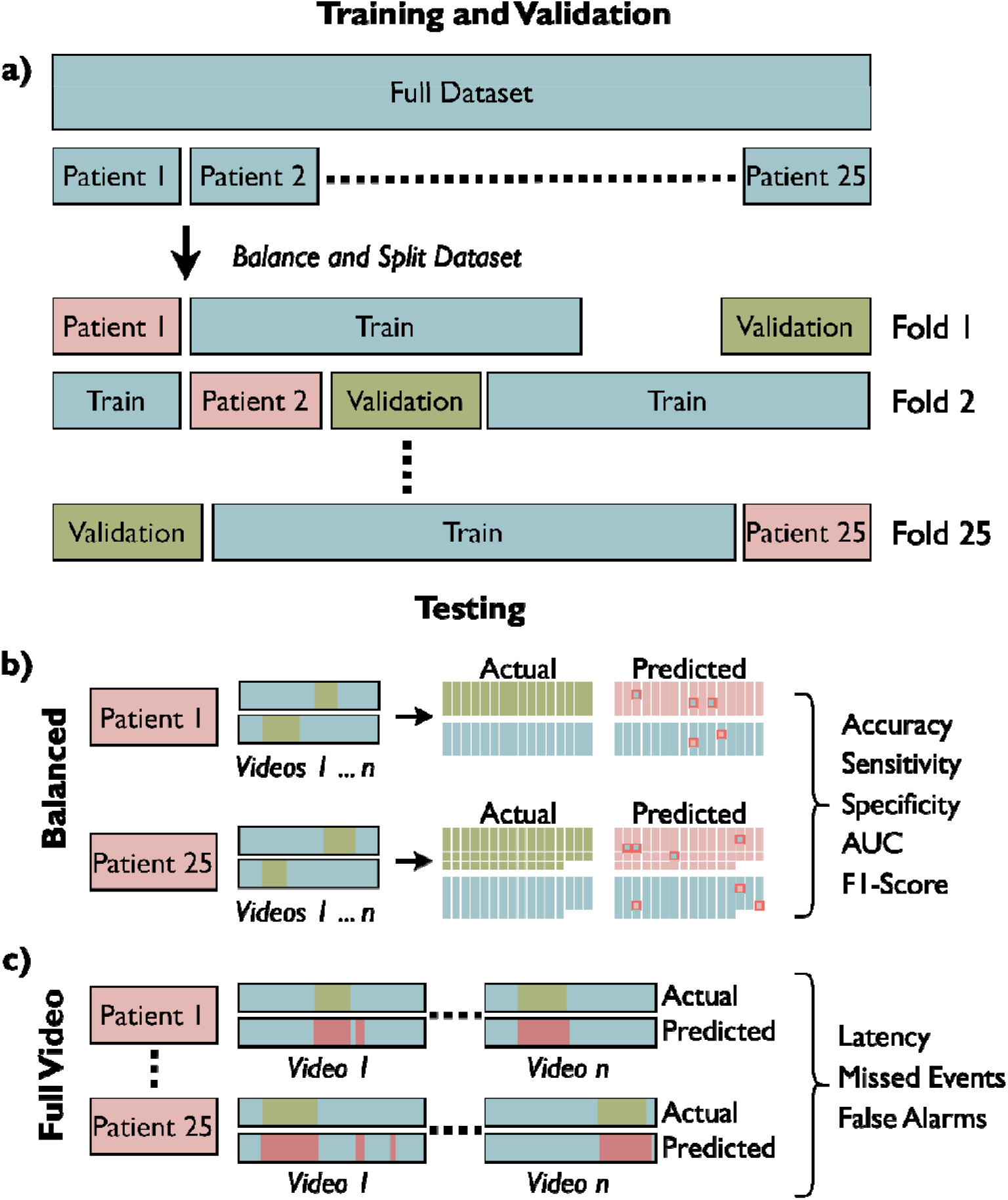
Model training and testing strategy. **a)** 25-fold, leave-one-patient-out cross-validation. In each fold, all videos from a single patient were set aside as the test set. The remaining clips were then selected to create a balanced dataset of seizure and non-seizure classes as described in section 2.4 then split into training and validation sets at a ratio of 80:20. **b)** Balanced testing. All videos from the test patient were divided into clips then balanced as described in section 2.4. The model was then used to classify the individual clips as seizure or non-seizure to derive model accuracy, sensitivity, specificity, area under the receiver-operator curve (AUC) score, and f1-score. **c)** Full video testing (unbalanced). Model evaluation was repeated using all video clips from the corresponding test patient in sequential order to approximate use in a clinical scenario and evaluate detection latency, missed seizure events, and false alarms.

The balanced test set consisted of all seizure video clips and an equal number of randomly selected non-seizure video clips from the patient left out of training. Training and evaluation using a balanced dataset ensured that the performance metrics were not skewed by bias to a non-seizure majority class. The unbalanced test set consisted of all video clips from the patient left out of training in sequence. Testing using sequential, unbalanced data was used to approximate the detection latency and false alarm rate, and to estimate model performance in an EMU setting where non-seizure activity is vastly more common than seizure.

### 2.5 Cross-Institutional Generalization, Training and Evaluation

After training and testing at the initial site (site A), the code for model training and testing was sent to the University of California, Los Angeles (site B). Site B then trained and cross-validated another series of seizure detection models on a separate dataset collected at their institution. This segment of the study was independently approved by the Institutional Review Board at the University of California, Los Angeles. All site B videos were recorded using a SONY EP580 camera at an original resolution of 1080×1920 pixels and comprised 3.13 total hours of EMU video data with 22.7**%** of the video data exhibiting seizure activity as identified by an epileptologist (cohort characteristics further elaborated in **Supplementary Table 1**). All videos were downscaled to match the 640×480 pixel resolution at site A. The model structure, initial pre-trained weights, and cross-validation process were identical between the two sites. These results were used to test the generalizability of the underlying model architecture and training process.

After evaluating the site B models on the site B dataset, the model with the best overall performance from site A was evaluated on the full dataset from site B. Likewise, the best performing model from site B was evaluated on the full dataset from site A. This process was designed to test the generalizability of a fully trained model to a dataset with different environmental conditions such as camera type, camera location, and bed setup.

### 2.6 Measures

Performance of the model during cross validation was evaluated using accuracy, area under the receiver operator curve (AUC) score, sensitivity, specificity, and F1-score (14,20). These metrics were computed on a balanced dataset by using a random subset of non-seizure clips equal to the number of seizure clips. Equations for performance measures are given in **Supplementary Figure 1**. Cross-validated model performance was evaluated on the full test sets videos, which are unbalanced, using detection latency, false alarm rate, and missed events. These metrics are based on the weighted moving median (WMM) of the classifier output from 3 second clips, an aggregate of model predictions over time defined in **Supplementary Figure 2**. A true alarm is defined as a WMM greater than 0.5 during the seizure or in the 20 seconds immediately preceding a patient seizure. Conversely, a false alarm is defined as a WMM greater than 0.5 at any other time during the video, with a 60 second buffer where no false alarms can be triggered after a true or false alarm. The false alarm rate is reported as average false alarms per hour, averaged over all videos in the test set. The percentage of full videos in each test set with zero false alarms is also reported. Detection latency is defined by the time in seconds from the start of a seizure to a true alarm with a minimum value of 0 seconds. A missed event is defined as a seizure where the model fails to sound a true alarm (**Supplementary Figure 2**).

## 3. RESULTS

### 3.1 Model Performance on the Balanced Dataset

Individual metrics for each patient evaluated in LOPO cross-validation are available in **Supplementary File 1**. Average accuracy was 95.94% ± 0.75% and average AUC was 0.988 ± 0.004. These results represent classification performance of the model on isolated 3-second video clips, with an even balance of seizure and non-seizure clips. The full results of balanced testing are presented in comparison to the prior best-in-class model in **Table 2**.

**Table 2:**
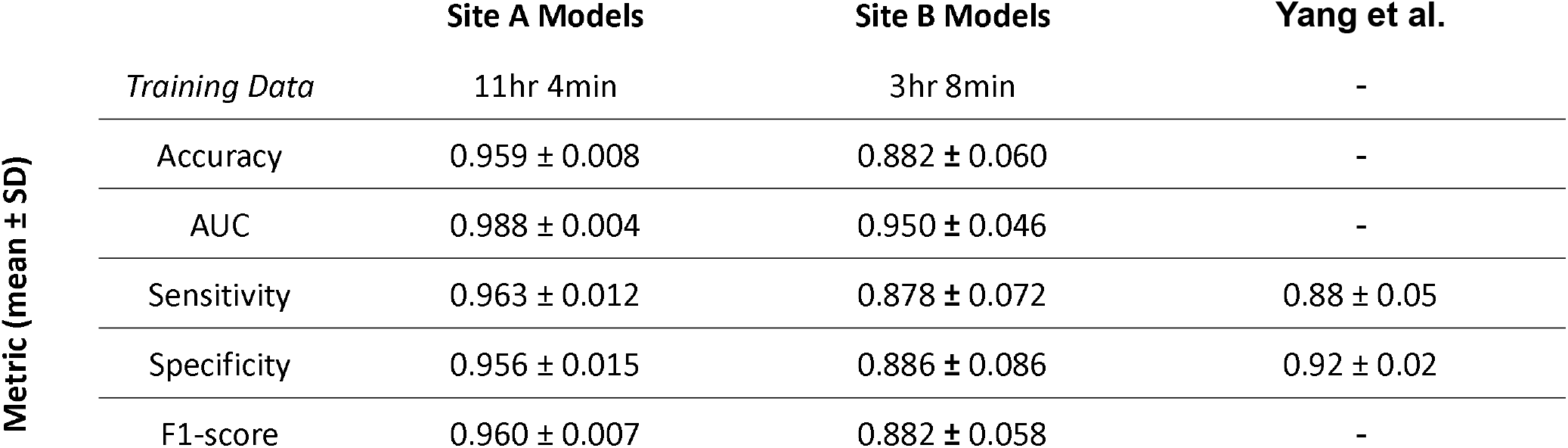
Summary of test evaluation metrics achieved by models at sites A and B during balanced testing with comparison to prior best in class 2D-CNN Long Short Term Memory (LSTM) model by Yang et al. (10). (Abbreviations: SD, standard deviation; hr, hours; min, minutes)

### 3.2 Full-Video Classification Performance

The models from each cross validation fold were also used to sequentially classify all video clips of the patient left out in the corresponding test set without balancing the data to better emulate clinical use. In these full video tests, all seizures were successfully detected with a median detection latency of 0.0 seconds IQR 0.0-3.0 seconds). Thirty-six of the forty-nine (73.5%) videos classified had no false alarms. Across all videos, the average false alarm rate was 1.81 per hour. False alarm rate did not vary significantly with video length (p = 0.102) as measured by Spearman’s rank correlation coefficient. Summary statistics for model performance across the recordings are shown in **Table 3** and plots for each video are available in **Supplementary Figure 3**.

**Table 3:** Summary of test evaluation metrics achieved by the models at both sites when classifying full, previously unseen videos at the same site as training (bolded) and at the other site. Abbreviations: IQR, interquartile range; s, second; hr, hour; BCI, 95% binomial exact confidence interval computed via the Clopper-Pearson method.

|  | Site A Models |  | Site B Models |  |
| --- | --- | --- | --- | --- |
|  | Site A Data | Site B Data | Site A Data | Site B Data |
| Detection Latency (median, IQR) | **0.0s (0.0, 3.0)** | 19.5s (4.5, 42.0) | 33.0s (22.5, 42.0) | **6s (3.0, 11.25)** |
| Mean False Alarm Rate | **1.81/hr** | 5.43/hr | 0.63/hr | **0.64/hr** |
| False alarm free videos (% , BCI) | **73.5% (61.2-85.7%)** | 57.9% (36.8-78.9%) | 89.8% (81.6-98.0%) | **94.7% (81.8-100%)** |
| Missed Seizure Events (% , BCI) | **0.0% (0.0-5.9%)** | 5.3% (0.0-15.8%) | 28.6% (16.3-40.8%) | **5.3% (0.0-15.8%)** |

**Figure 3:**
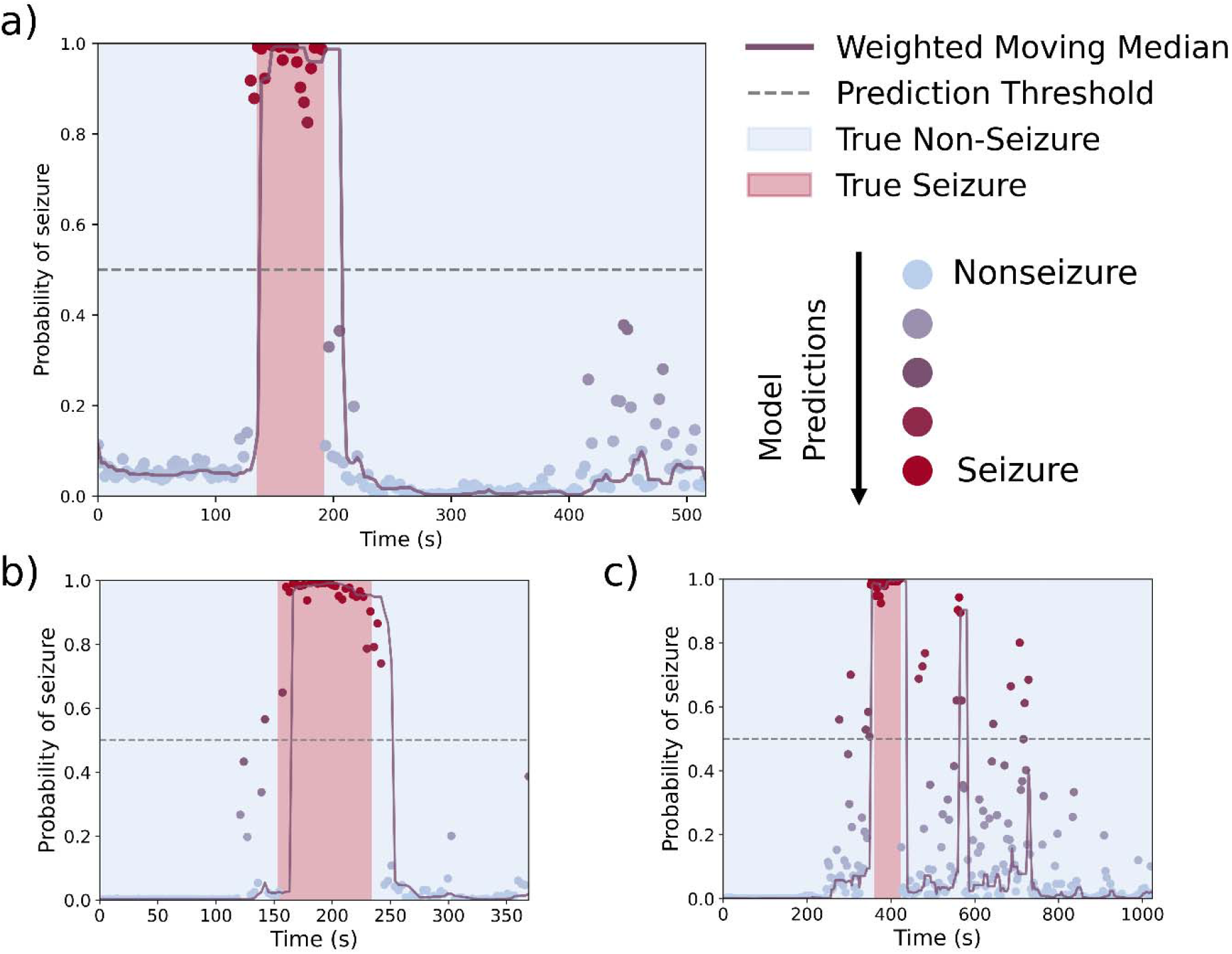
Visualization of model predictions and associated metrics on full videos. Individual model predictions (colored dots) are aggregated using the weighted moving median (lavender line). An alarm occurs when the weighted moving median crosses the prediction threshold of 0.5 (grey dotted line). True alarms occur during a true seizure, indicated by the red background color, or in the immediate preictal (20 seconds before seizure) or postictal (60 seconds after seizure) period. False alarms occur at any other time, with a buffer of 60 seconds before another alarm can be triggered. **a)** Video segment with low (3 second) detection latency, no missed events, and no false alarms. **b)** Example of a video with a relatively long (12 second) detection latency but no false alarm as the moving average returns to baseline within 60 seconds of seizure end. **c)** Video with model uncertainty and a false alarm during the post-ictal period. Plots for all videos at sites A and B can be found in **Supplementary Figure 3**. Abbreviations: s, seconds.

### 3.3 Generalizability of Architecture and Training Scheme

To validate the model architecture and training scheme on an independent data set, code for training and LOPO cross-validation were sent to site B. The results from the balanced testing are shown in **Table 2**. The site B model obtained a lower f1-score than the site A model (0.882 vs. 0.960, p < 0.005) as determined by the one-sided Kolmogorov-Smirnov test. Results from full video testing of the site B model on the corresponding 19 videos EMU recordings are shown in **Table 3**.

### 3.4 Cross-Institutional Testing

To investigate the generalizability of already trained models on novel datasets with different environmental conditions, the model with the best performance during balanced testing at site A was sent to site B and used to classify the 19 EMU videos from site B. Likewise, the best performing model from site B was sent to classify the 49 EMU videos from site A. Both models performed worse during cross-institutional testing with a significant increase in detection latency when compared with same-site performance (p < 0.005) as determined by the one-sided Mann-Whitney U test. A summary of the results is shown in **Table 3**.

### 3.5 Performance

The model runs at approximately 1,800 frames per second on a PC using a single GeFORCE RTX 4090 graphics card.

## 4. DISCUSSION

Our results demonstrate that video-only seizure detection using a spatiotemporal 3-D CNN model is feasible and highly accurate. The I3D architecture, initially developed for human action recognition, is highly effective in detecting tonic-clonic seizures using only video data. Furthermore, our results show the efficacy of using transfer learning between different clinical sites to fine-tune a large, pretrained activity classification model for clinical tasks with only a modest sized dataset. This finding builds upon previous work showing that video-based methods can robustly capture and analyze epileptic seizures (14).

To the best of our knowledge, our model’s performance for detecting tonic-clonic seizures exceeds all reported video-only methods, including the prior best-in-class 2D-CNN LSTM model developed by Yang et al. (10) and the pipeline incorporating an I3D model as a feature extractor but not performing fine-tuning of the backbone by Karácsony et al. (9). Like Yang et al., we employed LOPO cross-validation for training and balanced testing, a method shown to be a more reliable estimator of model performance compared to k-fold cross-validation that does not respect patient boundaries (21). This approach ensures that the model’s performance metrics are not inflated by data leakage and provides a more realistic assessment of the model’s generalizability to new patients.

With respect to full video testing, the low detection latency and absence of missed seizure events in full video testing at site A highlight the model’s reliability in a simulated clinical setting, although the frequent false alarms pose a challenge to implementation. Importantly, most false alarms occurred in a small subset of videos (73.5% contained no false alarms) suggesting that this obstacle may be addressable with further model tuning. However, it should be noted that videos were obtained retrospectively. As such, the longest video tested was only 33 minutes, and future testing with longer recordings is necessary to comprehensively evaluate the false alarm rate.

An important contribution of this work is the demonstration of the generalizability of the model architecture across different clinical environments. Despite training on significantly less data, the model architecture and training achieved relatively high performance when used to train and evaluate models at site B. This finding suggests that an I3D model in conjunction with our training pipeline can be effectively deployed across different EMUs if provided limited site-specific training data. This aspect of our study is crucial for practical implementation, as it implies that the model can be scaled and adapted to new clinical settings with minimal effort, a key consideration for widespread adoption (22).

Though the underlying architecture transferred well between institutions (**Table 2**), the model trained at site A failed to generalize to site B data and vice versa (**Table 3**). In particular, the significantly higher detection latencies and greater number of missed seizure events suggest that models were not able to fully generalize to new features in the EMU environment not seen at the other institution. Nonetheless, the models performed significantly better than chance, and it remains unclear if more training data or multi-institutional datasets could close the gap between same-institution and cross-institution performance. Federated learning, a technique designed to allow for model training on multi-institutional data without the need to centralize videos in one location, is a promising solution to address legal and privacy concerns associated with sharing protected health information and facilitate multi-institutional collaboration (23). Though the current model pipeline does not include features for real-time detection from streamed video data, the high video processing speed exceeding 1,800 frames per second suggests that real-time detection from one or more video feeds could be possible. Moreover, the lack of pre-processing requirements, such as cropping or other modifications to the video data, minimizes variations between institutions and reduces processing time, further enhancing the model’s potential applicability in real-time scenarios such as the EMU or home seizure monitoring.

This work focused only on tonic-clonic seizure detection, and it is likely that the model would not perform as well for seizures with limited movement such as purely tonic, absence, or focal without progression to tonic-clonic. Future work including a broader array of seizure types during training could potentially address this limitation in our model. Training and validation on home seizure detection setups with variable layout and quality is also an essential next step for a truly general-purpose model. Other areas for future work include standardization of clinical metrics such as detection latency, tests with full 24 hour videos to better characterize false alarm rate, and clinical trials for the implementation and evaluation of real-time, I3D model based seizure detection in the EMU (20). Integration of audio data into model designs, as done by several existing investigational and commercial seizure detection algorithms, may also increase overall model performance and generalizability (24).

## 5. CONCLUSION

Our study underscores the feasibility of using a spatiotemporal 3-D CNN model for video-only detection of tonic-clonic seizures with high accuracy and speed. The model’s high processing speed, minimal pre-processing requirements, and robust performance across different clinical environments when provided sufficient training data make it a potentially valuable tool for seizure monitoring. Future enhancements, including larger and more varied datasets, the integration of audio data, and the use of federated learning, can further augment the model’s capabilities and extend its applicability in diverse clinical settings. By leveraging advanced computer vision techniques, we can significantly improve the accuracy and efficiency of seizure detection, ultimately enhancing patient care and clinical outcomes in epilepsy management.

## Supporting information

Complete Results

## Data Availability

Data Availability Statement: Code used to generate the results in this study is based on the MMaction2 toolbox and is publicly available at https://github.com/aidanboyne/CVEpilepsy under the Apache 2.0 license. The video data is not available due to patient privacy issues.

## Acknowledgments

None.

