## Supplementary Figures for "VIDEO BASED DETECTION OF EPILEPTIC SEIZURES USING A THREE-DIMENSIONAL CONVOLUTIONAL NEURAL NETWORK"

| ID | Seizure length (s) | Seizure % | Non-SeizuRe length (s) | Non-Seizure % | Total length (s) |
| --- | --- | --- | --- | --- | --- |
| 00001 | 72.3 | 12% | 554.7 | 88% | 627 |
| 00002 | 61.8 | 8% | 669.2 | 92% | 731 |
| 00003 | 125.7 | 20% | 493.9 | 80% | 619.6 |
| 00004 | 76.8 | 17% | 374.2 | 83% | 451 |
| 00005 | 101 | 19% | 426 | 81% | 527 |
| 00006 | 70 | 19% | 305.9 | 81% | 375.9 |
| 00007 | 70.3 | 13% | 478.6 | 87% | 548.9 |
| 00008 | 75.5 | 22% | 270.4 | 78% | 345.9 |
| 00009 | 107.3 | 22% | 371.7 | 78% | 479 |
| 00010 | 66.4 | 12% | 479.6 | 88% | 546 |
| 00011 | 130.1 | 24% | 413.9 | 76% | 544 |
| 00012 | 180.1 | 40% | 270.9 | 60% | 451 |
| 00013 | 115.1 | 19% | 475.9 | 81% | 591 |
| 00014 | 118.5 | 24% | 375.5 | 76% | 494 |
| 00015 | 393.9 | 43% | 523 | 57% | 916.9 |
| 00016 | 103.2 | 18% | 470.8 | 82% | 574 |
| 00017 | 85.8 | 17% | 410.2 | 83% | 496 |
| 00019 | 282.7 | 21% | 1035 | 79% | 1317.7 |
| 00020 | 315.8 | 51% | 300.2 | 49% | 616 |

**Supplementary Table 1**: Breakdown of video length and composition by patient at site B.

**Supplementary File 1:** For summarized output of all testing and training for models at sites A and B, see *All_Results.xlsx*.

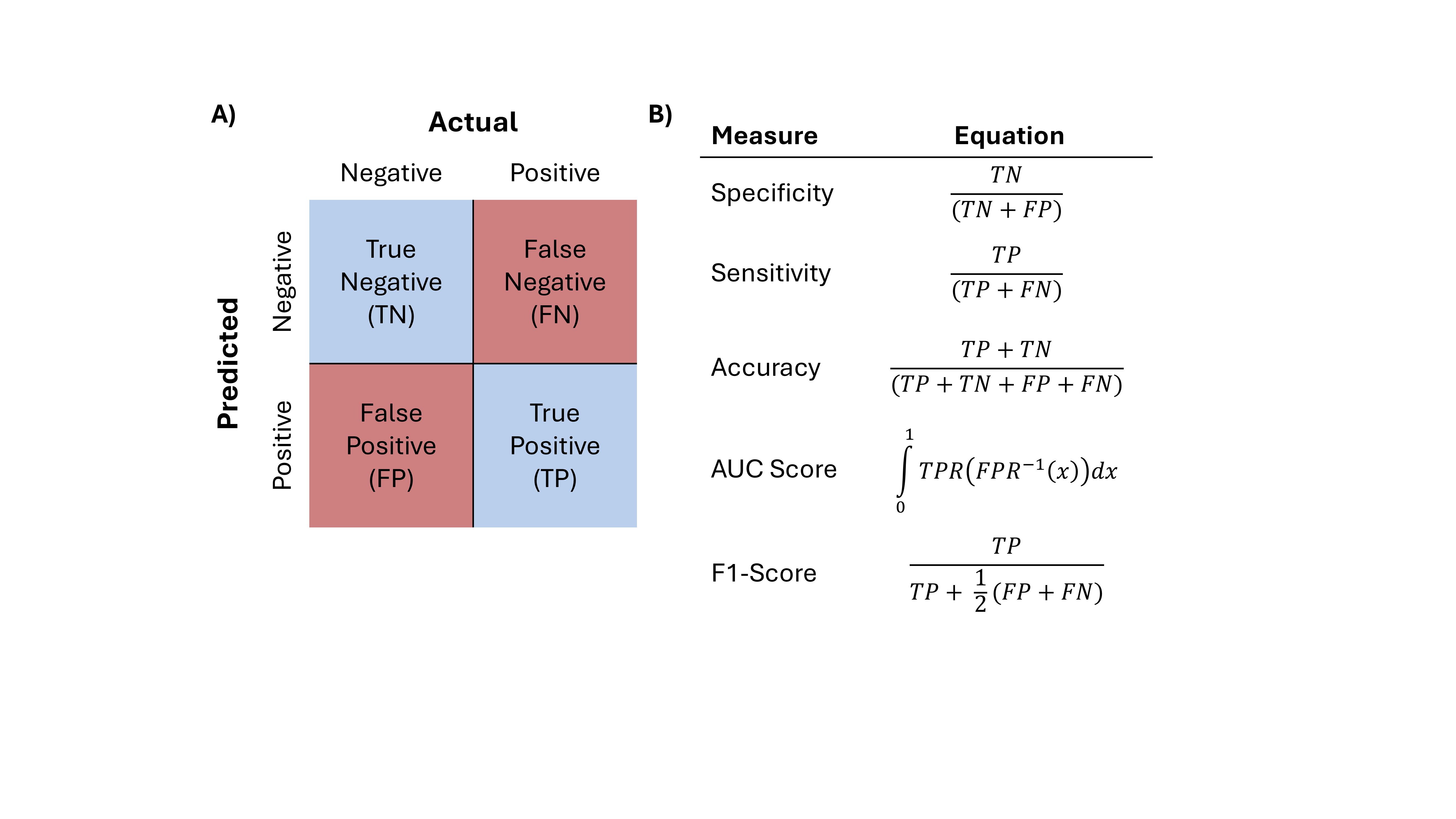

**Supplementary Figure 1:** Mathematical definitions of statistical measures used for model evaluation. All metrics range between 0 and 1, where 1 represents the best possible performance. (A) Video clips are predicted as seizure (positive) or non-seizure (negative) by the model then compared to the actual label determined by an epileptologist to create a confusion matrix. (B) Entries from the confusion matrix are used to calculate specificity, sensitivity, and accuracy. Area Under the Receiver Operating Curve (AUC Score) is calculated by finding the area under a curve generated by varying cut-offs for sensitivity vs. 1-specificity. AUC score provides a balanced measure of model performance relatively robust to imbalanced data such as full EMU videos where non-seizure activity is more common than seizure.

**a)**

**Algorithm -** Weighted Moving Median

**Require:** data, *k*, weights = [*w*_1_*, w*_2_*, w*_3_*, w*_4_], cutoffs = [*c*_1_*, c*_2_*, c*_3_*, c*_4_] **Ensure:** Weighted moving median values *M* = *{m*_1_*, m*_2_*, . . . , m_n_}*

1: Initialize an empty deque window with maximum length *k*.

2: Initialize an empty list *M* to store the results.

3: **for** each *x ∈* data **do**

4:	Compute weight *w* for *x* based on cutoff thresholds cutoffs.

5:	Append (*x, w*) to window.

6:	**if** window is full **then**

7:	Extract values *x*_i_ and weights *w*_i_ from window.

8:	Sort values and weights by ascending values.

9:	Compute cumulative weights cumsum(*w*_i_).

10:	Find the smallest index *i* such that cumsum*_i_ ≥* 0*.*5 *×* total weight.

11:	Assign *m ← x*_i_.

12:	Append *m* to *M* .

13:	**else**

14:	Append None or NaN to *M* (optional for incomplete windows).

15:	**end if**

16: **end for**

17: **return** *M*

**c)**

**b)**

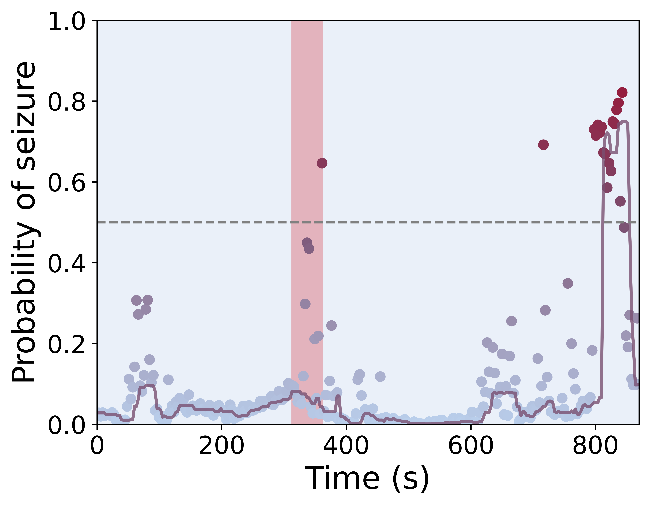

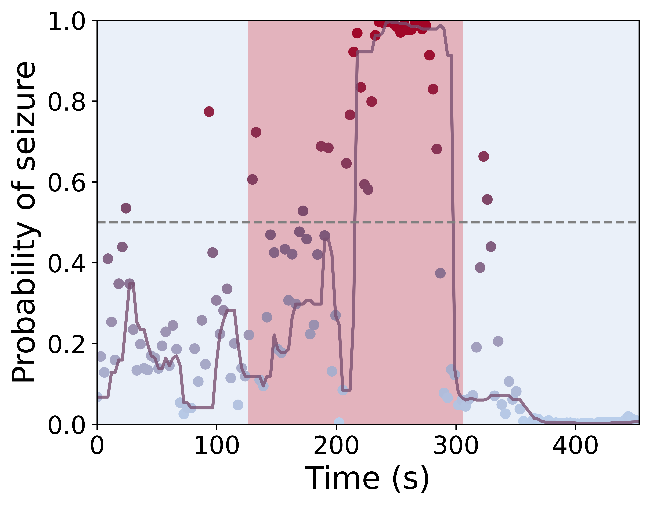

**Supplementary Figure 2**: Definitions for full video test metrics. **a)** Weighted moving median algorithm. The implementation in the code uses k = 8, weights = [3, 1, 0.5, 1, 5] and cutoffs = [0.1, 0.35, 0.65, 0.9] to prioritize more confident predictions while penalizing uncertain ones, giving extra weight to highly confident seizure predictions in an attempt to prevent missed events. Weights, cutoffs, and window size *k* were chosen by hand for interpretability and could likely be optimized in a clinical scenario. **b**) Plot depicting a missed seizure, indicated by the failure of the weighted moving median (lavender line) to cross the prediction threshold (grey dotted line) during the seizure (red background), and a false alarm. **c)** Plot with high detection latency of 90 seconds, indicated by the thick red bar overlaid on the plot.

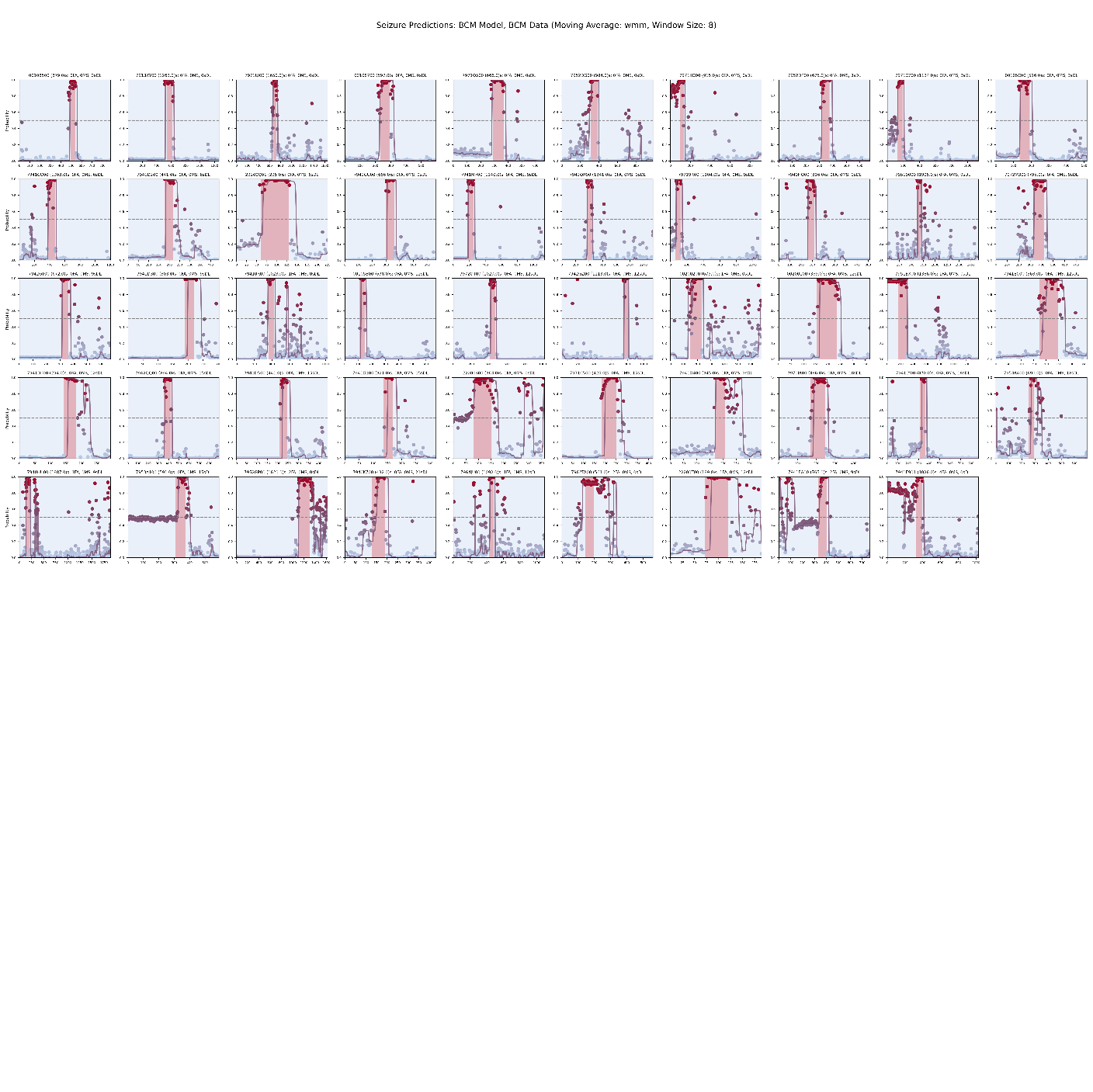

**b)**

**a)**

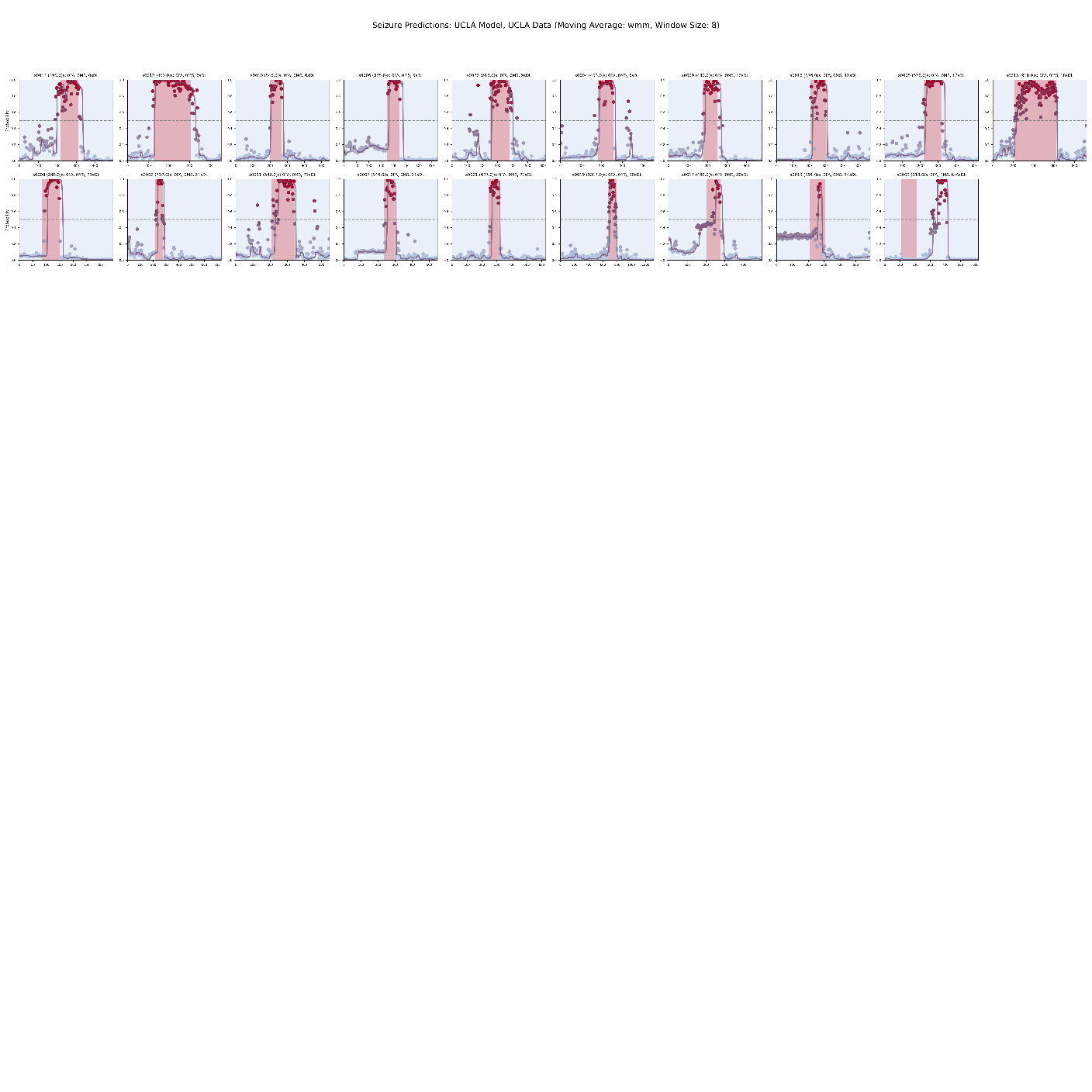

**c)**

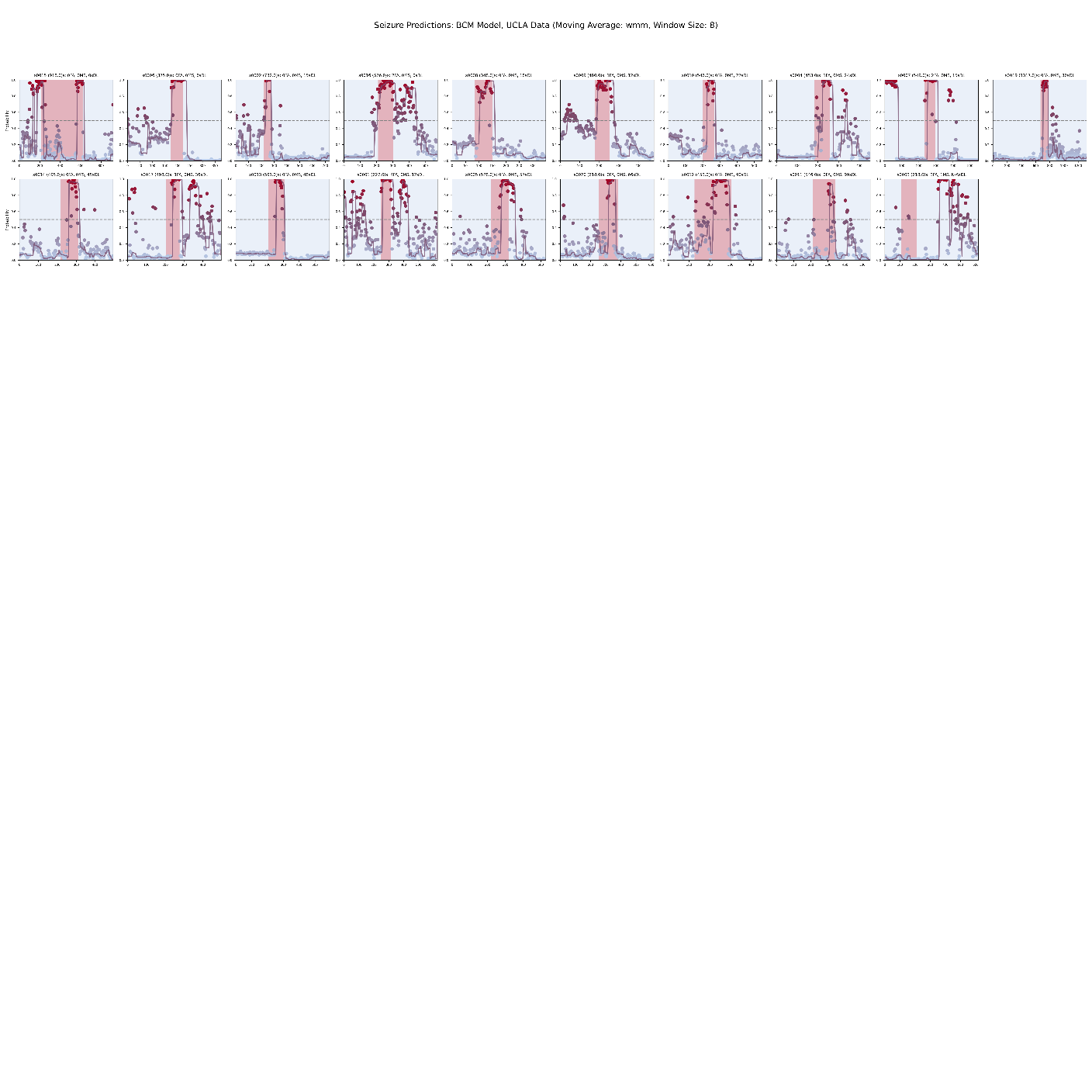

**d)**

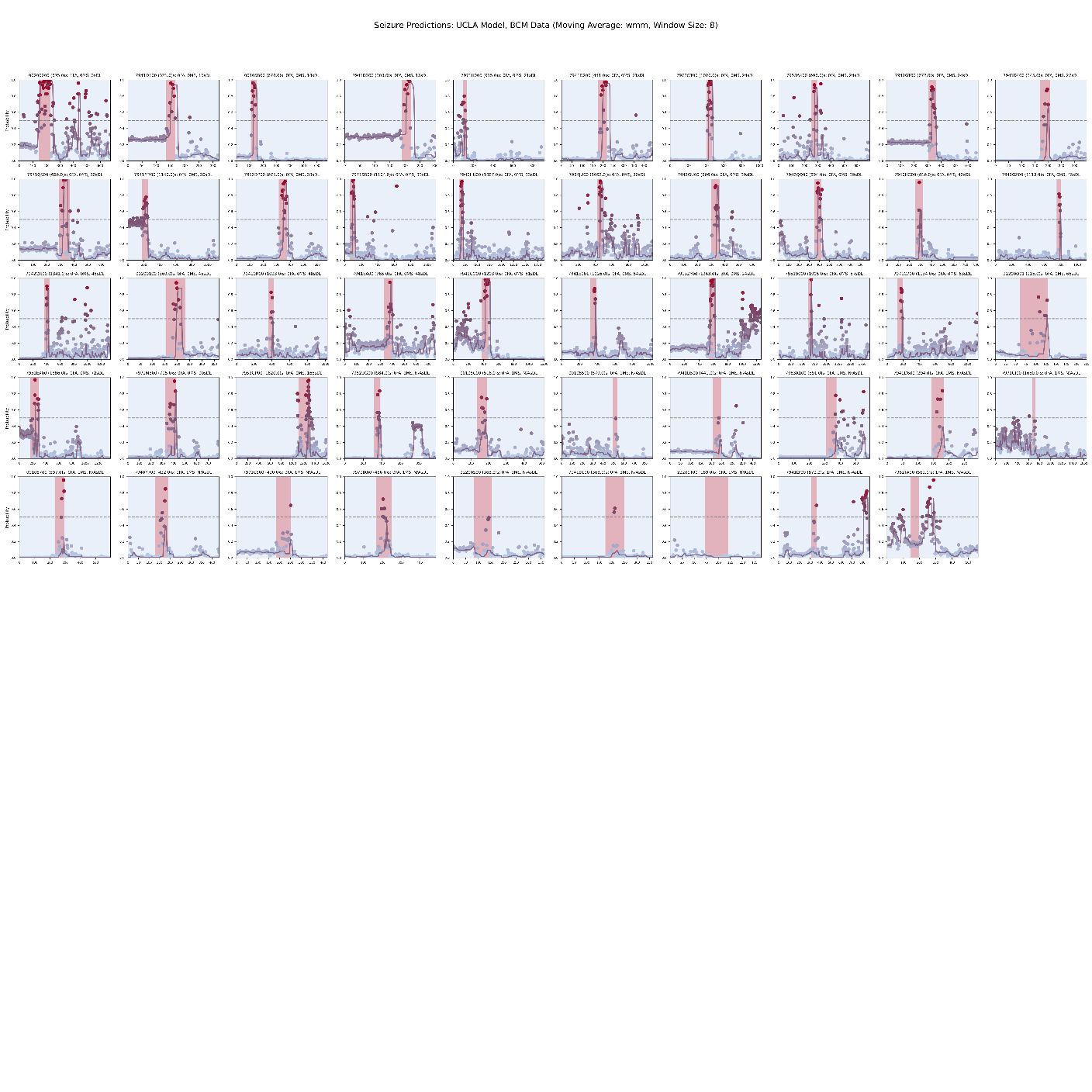

**Supplementary Figure 3**: Visualization of model predictions and associated metrics on full videos. Individual model predictions (colored dot) are aggregated using the weighted moving median (lavender line) and overlaid on the corresponding segment of true non-seizure (light blue background) or seizure (red background) activity. Movement of the weighted moving median above the prediction threshold (grey dotted line) indicates an alarm. For each site and dataset, plots are roughly ordered from best to worst performance based on missed events, false alarms, and detection latency. **a)** Full video, same-site testing of the site A model on site A videos. **b)** Full video, same-site testing of the site B model on site B videos. **c)** Full video, cross-site testing of the site A model on site B videos. **d)** Full video, cross-site testing of the site B model on site A videos. Abbreviations: s, seconds; FA, False Alarm(s); MS, Missed Seizure(s); DL, Detection Latency.

Full size plots for each subplot a-d are available as SuppF3_A.png, SuppF3_B.png, SuppF3_C.png, and SuppF3_D.png respectively.
