## Supplementary figures and images for "VIDEO BASED DETECTION OF EPILEPTIC SEIZURES USING A THREE-DIMENSIONAL CONVOLUTIONAL NEURAL NETWORK"

### Supplementary Figure 3a

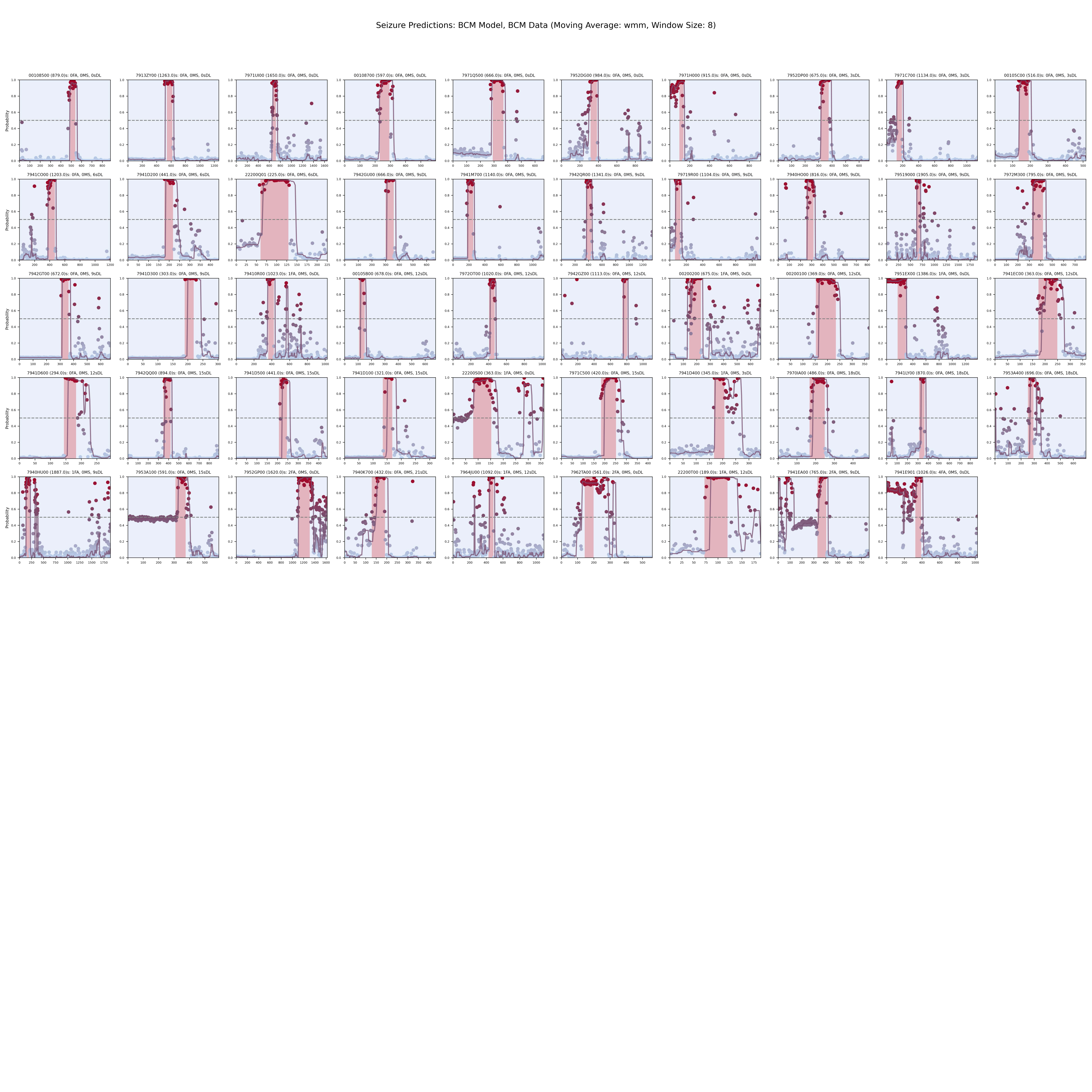

### Supplementary Figure 3b

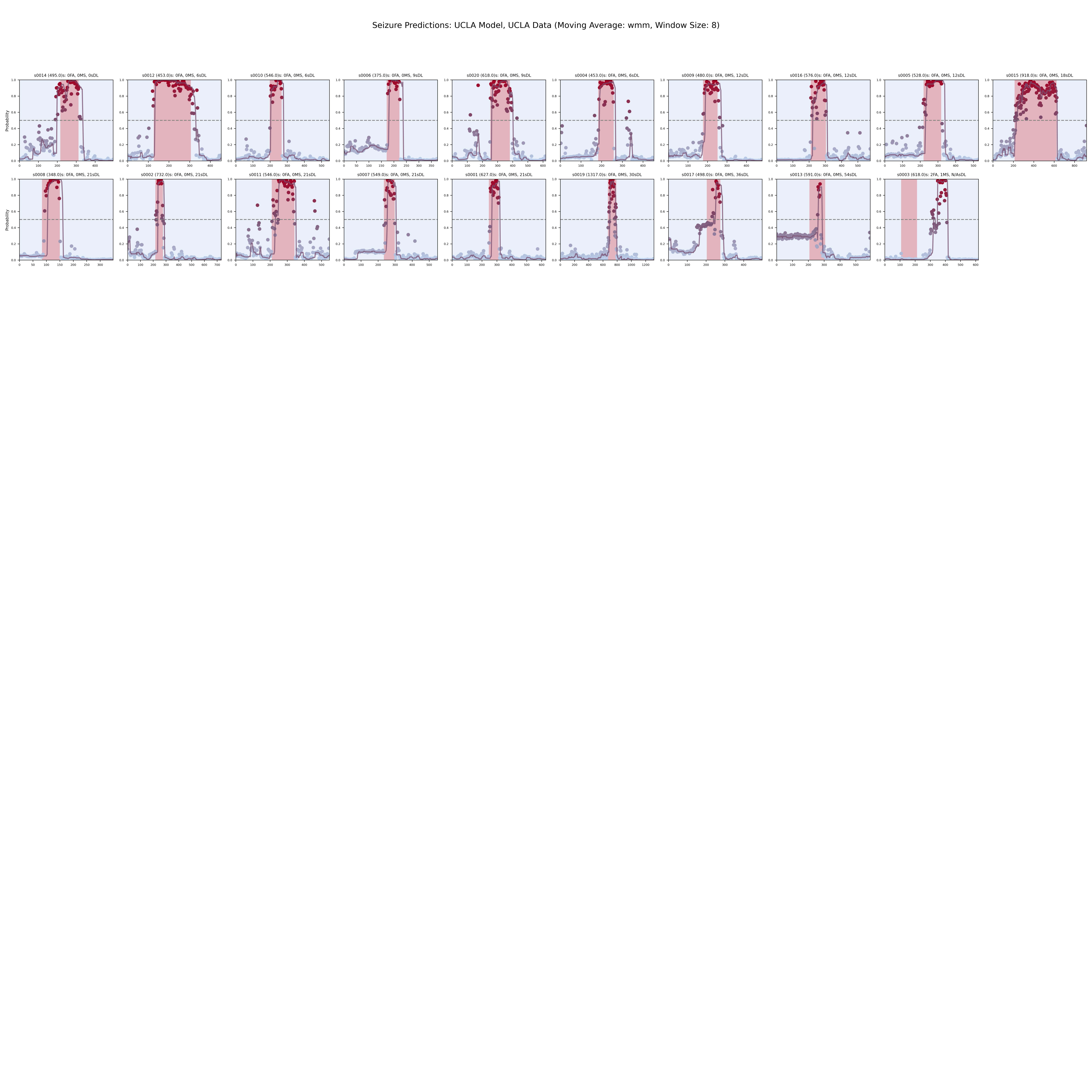

### Supplementary Figure 3c

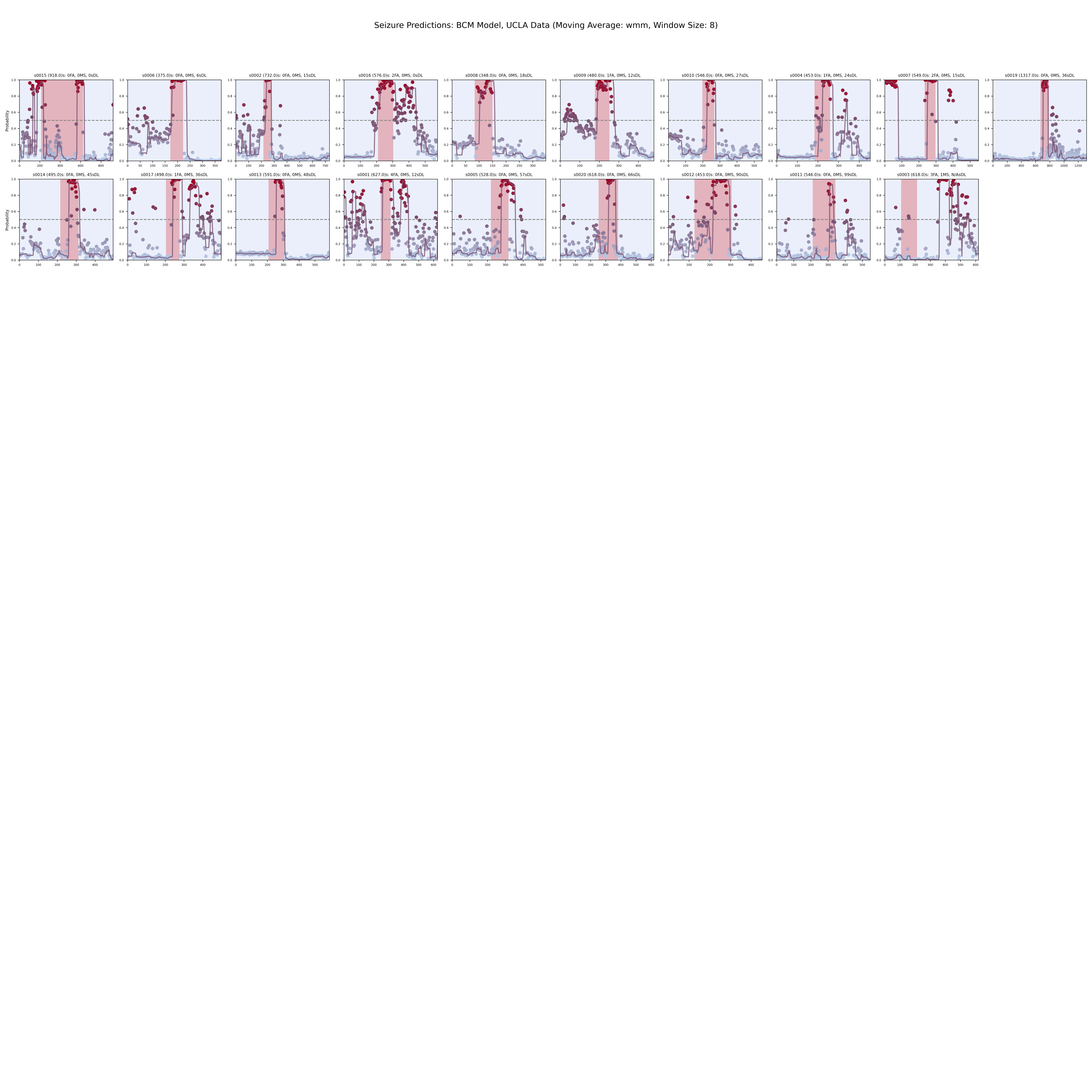

### Supplementary Figure 3d

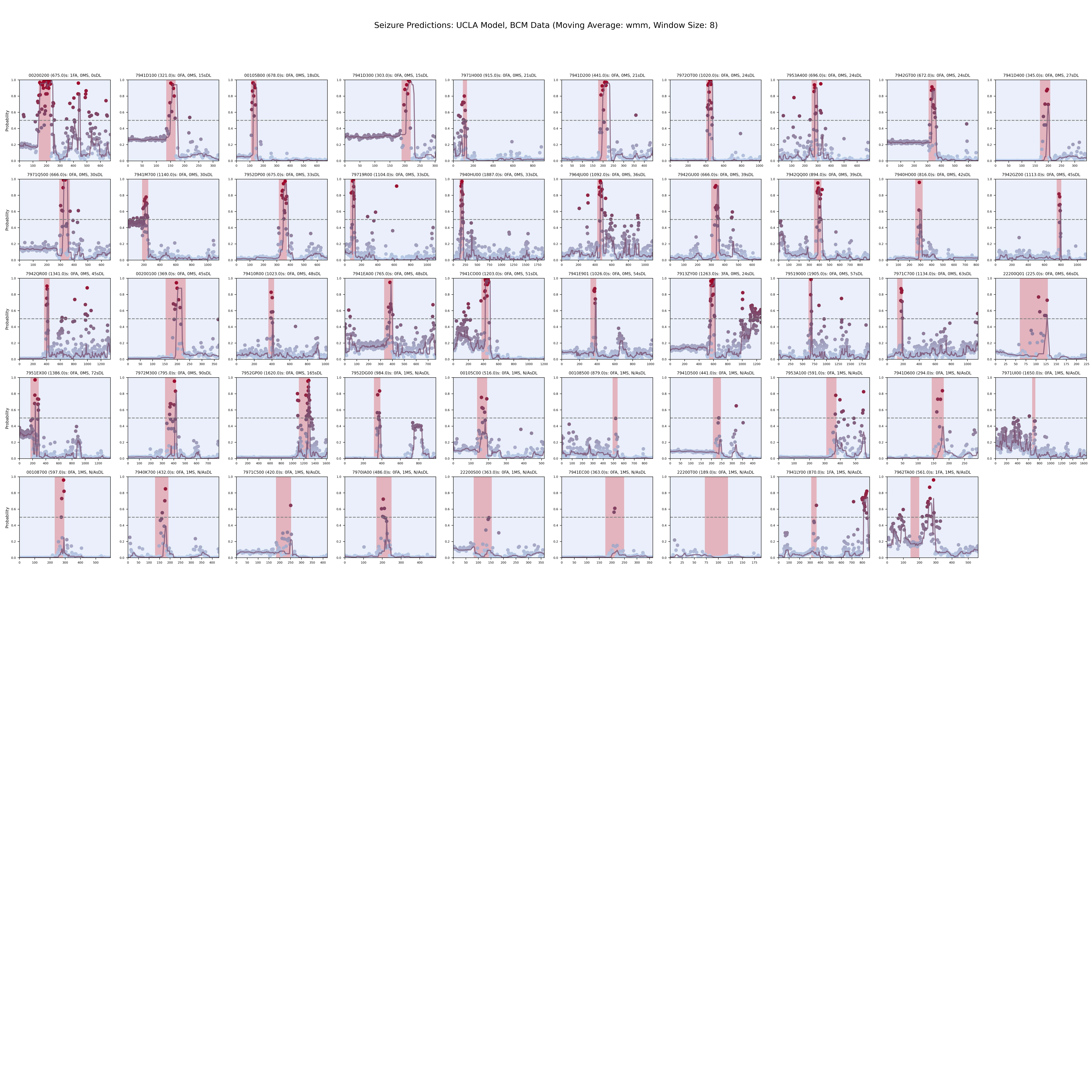
